# Effect of evidence updates on key determinants of measles vaccination impact: a DynaMICE modelling study in ten high-burden countries

**DOI:** 10.1101/2021.05.28.21257964

**Authors:** Han Fu, Kaja Abbas, Petra Klepac, Kevin van Zandvoort, Hira Tanvir, Allison Portnoy, Mark Jit

**Author notes:** **Corresponding author** Han Fu, PhD, London School of Hygiene & Tropical Medicine, Keppel Street, London WC1E 7HT, United Kingdom.

## Abstract

**Background:** Model-based estimates of measles burden and the impact of measles-containing vaccine (MCV) are crucial for global health priority setting. Recently, evidence from systematic reviews and database analyses have improved our understanding of key determinants of measles vaccine impact. We explore how updated representations of these determinants affect model-based estimation of MCV impact in ten countries with highest measles burden.

**Methods:** Using Dynamic Measles Immunisation Calculation Engine (DynaMICE), an age-structured compartmental model of measles transmission and vaccination, we evaluated the effect of evidence updates for five determinants of MCV impact: case fatality risk, contact patterns, age-dependent vaccine efficacy, the potential of supplementary immunisation activities (SIAs) to reach zero-dose children, and the basic reproduction number. We also evaluated the incremental impact of the first dose (MCV1), second dose (MCV2), and SIA dose of measles vaccines, based on country-specific coverage estimates from the World Health Organization. The MCV impact was assessed by cumulative vaccine-averted cases, deaths, and disability-adjusted life years over 2000–2050.

**Results:** Incorporated with the updated data sources, DynaMICE projected 252 million measles cases, 3.7 million deaths and 230 million disability-adjusted life years incurred over 2000–2050 in the ten high-burden countries when MCV1, MCV2, and SIA doses were implemented. Compared to no vaccination, the administration of MCV1 contributed to 66% reduction in cumulative measles burden, while MCV2 and SIAs reduced this further to 89%. With routine and supplementary vaccination, India and countries with high vaccination coverage could maintain measles incidence below 1 per million. Among the updated determinants, shifting from fixed to linearly-varying vaccine efficacy by age and from static to time-varying case fatality risks had the biggest effect on the model projections of MCV impact. While varying the basic reproduction number showed a limited effect on vaccine-averted burden, updates on the other four determinants together led to an overall reduction of MCV impact by 0.87–26.7%.

**Conclusions:** High coverage of measles vaccine through both routine and SIA delivery platforms are essential for achieving and maintaining low incidence in high-measles burden settings. Incorporating updated evidence particularly on vaccine efficacy and case fatality risk reduces estimates of the impact of vaccination slightly, but its overall impact remains considerable.

## Background

Measles is a highly contagious disease that may result in severe morbidity and mortality, particularly in young children and in settings with poor access to treatment. Vaccination is a safe and effective measure for measles prevention and control, as seen in high-income countries since its first licensure in 1961 (Strebel et al., 2018). The optimal age-range of the first dose of measles-containing vaccine (MCV1) depends on the local variation in burden, seasonality, and birth rate (Metcalf et al., 2011), with areas with high birth rates and low vaccination coverage favouring earlier vaccination. In settings with ongoing measles transmission, the World Health Organization (WHO) recommends delivering MCV1 to children of 9 months old, and following up with the second dose (MCV2) for children at 15-18 months old (WHO, 2017). In addition among countries with weak health systems, supplementary immunisation activities through vaccination campaigns are highly effective in protecting under-immunised and zero-dose children by closing immunity gaps and interrupting measles transmission (WHO, 2017). Vaccination contributes to the establishment of high levels of population immunity required for measles elimination, which is verified by the absence of endemic measles transmission for at least 36 months (WHO, 2013). In 2011, the Global Measles and Rubella Strategic Plan was set up with a goal to achieve measles elimination in at least five WHO regions by the end of 2020 (WHO, 2011). However, coverage of MCV1 has stagnated since 2010 in many countries, and has been set back in 2020 due to routine immunisation service disruptions and mass vaccination campaign suspensions caused by the COVID-19 pandemic. This has increased immunity gaps and the risk of measles outbreaks (Gaythorpe et al., 2021).

Strategic investments to improve measles vaccine coverage globally are partially informed by model-based estimates of measles burden and the impact of vaccination. A recent analysis conducted by the Vaccine Impact Modelling Consortium (VIMC) found that 57% of all vaccine-related mortality reduction was due to measles vaccination in 98 low-and middle-income countries (LMICs) between 2000–2019 (Li et al., 2021). However, such estimates are highly dependent on our knowledge of key determinants of measles incidence and mortality as well as vaccination impact. Over the past decade, there have been substantial advances to our knowledge of measles case-fatality risks (CFRs) (Portnoy et al., 2019), social contacts driving person-to-person infection transmission (Prem, Cook & Jit, 2017; Prem et al., 2020), age-related vaccine efficacy (Hughes et al., 2020), the ability of SIAs to reach undervaccinated populations (Portnoy et al., 2018) and measles basic reproduction numbers (Guerra et al., 2017). Nonetheless, how this additional evidence on epidemiological, behavioural and programmatic determinants affects estimates of vaccination impact has never been systematically explored.

In this study, we investigated the extent to which recent evidence updates about these determinants affects model-based estimates of measles burden and vaccination impact. To do this, we used the Dynamic Measles Immunization Calculation Engine (DynaMICE), a population-based dynamic transmission model of measles vaccination that has been used to inform VIMC’s estimates. We investigated the extent to which recent improvements in our understanding of the determinants of measles vaccination impact affects the estimates of overall measles disease burden, the impact of vaccination, and the development of effective measles vaccination strategies.

## Methods

### DynaMICE -Dynamic Measles Immunization Calculation Engine

DynaMICE is an age-structured compartmental transmission model designed to assess the impact of measles vaccination globally. Susceptible individuals become infected after effective contact with an infectious person, and remain immune once they recover from their infection. Infants are born with or without maternal antibodies, depending on the immunity of their mothers. The population is further divided according to their received number of measles containing vaccine doses (see Figure 1). The age structure is composed of weekly age classes for the first three years of age, with annual age classes thereafter up to 100 years. The force of infection is calculated by multiplying an age-dependent per-capita contact rate with the total number of infectious people in the population and the probability of transmission per contact. The latter is calculated by scaling the next-generation matrix to reach a target basic reproduction number (R_0_) of 16, assuming the average duration of infectiousness is 14 days. Annual seasonality of measles transmission was also incorporated into the model structure. Maternal immunity was assumed to last for an average of six months after birth, while individuals who recover from measles disease or acquire effective vaccine protection develop lifelong immunity. Measles deaths are calculated by applying an age-specific CFR to the incidence of cases. The model was coded in R and Fortran-95, and is available at https://github.com/lshtm-vimc/dynamice. Details of model equations and parameters are included in Appendix (Table S1 and Section S1) and also described in previously published studies (Verguet et al., 2015; Li et al., 2021).

**Figure 1:**
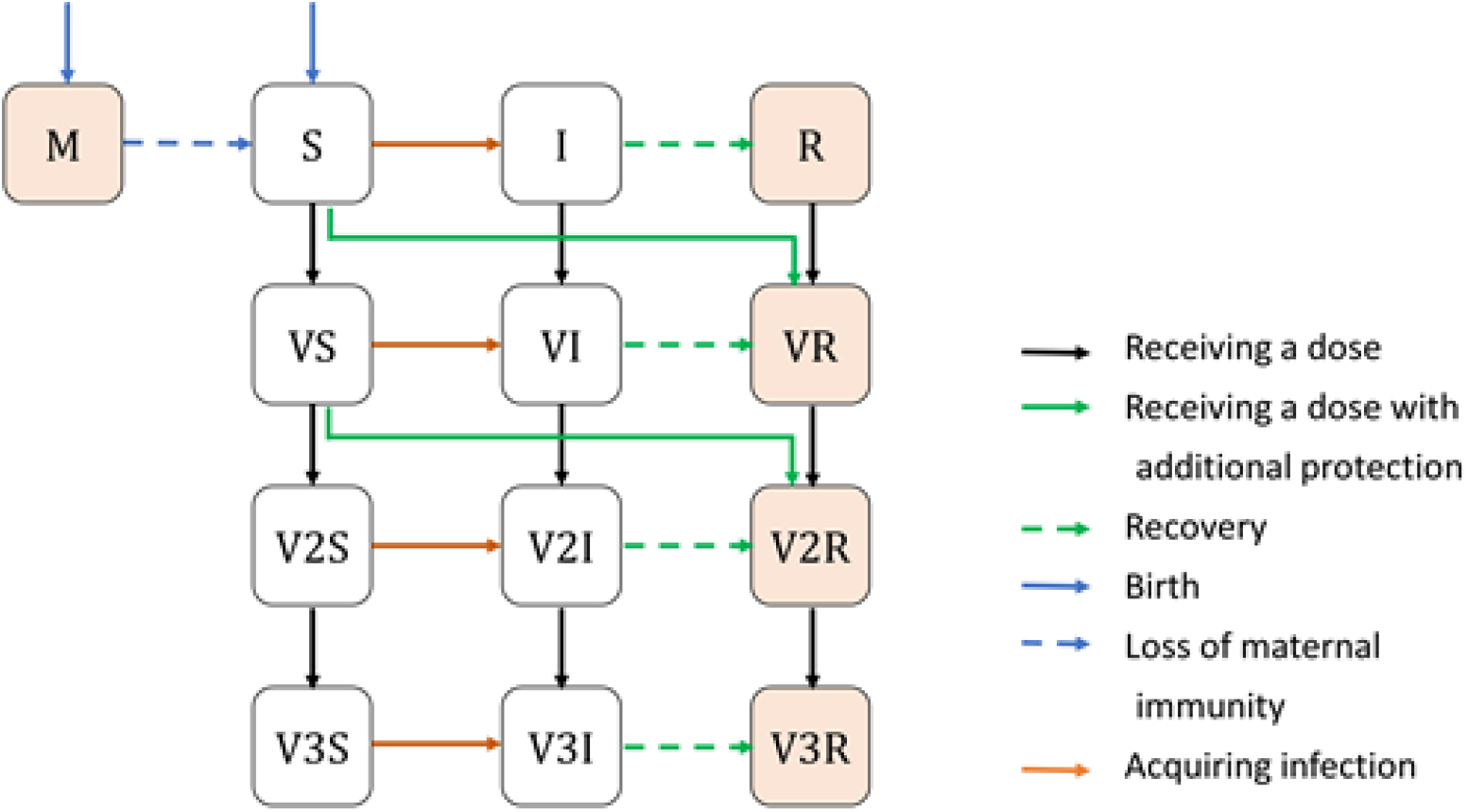
DynaMICE – Dynamic Measles Immunisation Calculation Engine model structure. Main model structure of DynaMICE (Dynamic Measles Immunisation Calculation Engine) for each age stratum. Individuals can be divided into 13 mutually exclusive states: S-susceptible, M-maternally immuned, I-infectious, R-recovered; V, V2, and V3 denotes the first, second, and third vaccine doses received. Individuals (orange shaded squares) become immune with natural or vaccine-acquired immunity against measles. Arrows denote the change between two states. In this model, births only add to the youngest age group. For clarity of presentation, ageing and death are not shown.

### Measles vaccination strategies

In the DynaMICE model, vaccination can be delivered to any age or range of ages and in either routine immunisation (MCV1, MCV2) or supplementary immunisation activities (SIAs). We assessed a range of model parameterisations from the perspective of four vaccination strategies: (1) no-vaccination, (2) MCV1 only, (3) MCV1 and MCV2, (4) MCV1, MCV2, and SIAs. We assumed that the implementation of MCV1 and MCV2 complies fully with the WHO-recommended schedule (WHO, 2017), delivered to children aged at 39 weeks (9 months) and 72 weeks (median of 15-18 months), respectively. MCV2 is delivered to children who have received one dose of measles vaccine. In addition to routine vaccination, SIAs target children of different age groups and occur at different intervals depending on settings. Historical coverage of vaccination was obtained from country-specific data in WHO databases (IVB/WHO, 2020a; WHO, 2020a,b), and we aggregated the number of populations reached by SIAs across subnational districts to obtain national coverages (Section S3 in Appendix). Future vaccine coverage for routine immunisation was projected to increase by 1% per year from 2019, up to a maximum of 95%, while SIAs were assumed to take place every three years after 2019, with the same coverage as the last recorded historical SIA (Figure S1). We modelled vaccine efficacy as all-or-nothing, that is, offering complete protection to a defined proportion of the vaccinated population, and no protection to the remainder of the vaccinated population. The efficacy for the first dose was assumed to depend on the age of vaccination (with further explanation in the later sections), while the combined first and second dose was assumed to have 98% efficacy (Sudfeld, Navar & Halsey, 2010). No additional protection was assumed to be received from any further doses of measles vaccine.

### Measles vaccination impact metrics

We modelled measles transmission and vaccination impact in ten highest measles burden countries in 2000 – India, Nigeria, Pakistan, Ethiopia, Afghanistan, Sudan, Tanzania, Niger, Somalia, and Democratic Republic of the Congo (DR Congo), together accounting for more than 65% of global measles mortality in that year (GBD 2019 Diseases and Injuries Collaborators, 2020). For each country and vaccination strategy, we projected time trends and age distributions of measles cases, deaths, and disability-adjusted life years (DALYs). We estimated the number of new infections, defined as incident measles cases, over 2000–2050. Measles-related deaths were calculated by multiplying the estimated number of measles cases with the specified age-specific CFR (Wolfson et al., 2009; Portnoy et al., 2019). DALYs for age group *a* at year *k* in country *c*were further calculated by:

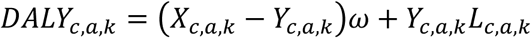

where *X*_*c,a,k*_ and *Y*_*c,a,k*_ represent number of cases and deaths, respectively; *ω* denotes the product of disability weight and length of illness, equal to 0.002 (Global Burden of Disease Collaborative Network, 2020); *L*_*c,a,k*_ denotes remaining life expectancy at age of death (United Nations, 2019). No time discounting was applied to the DALY calculation. We also assessed the calendar year for a country to have conditions suitable to achieve measles elimination, based on an approximate criterion — measles incidence has been maintained at <1 case per million population for at least three consecutive years (Wharton, 2004; WHO, 2013). In assessing the key determinants of measles vaccination impact, we calculated cumulative vaccine-averted burden over 2000–2050 by comparing the vaccination strategies to the baseline with no vaccination.

### Evaluation of determinants

We updated the data sources for five key determinants of vaccination impact in DynaMICE: case fatality risk, social contact patterns, age-dependent vaccine efficacy, proportion of zero-dose children reached by SIAs, and basic reproduction numbers (Figure 2). For the first four determinants, we first modelled a ‘base’ scenario based on previously adopted data sources (Verguet et al., 2015; Li et al., 2021) and then altered each determinant individually to reflect recent evidence updates. We also evaluated a ‘full-update’ scenario that included all the updated data sources for four of the determinants (Table 1), and examined the resulting changes in measles vaccination impact. For the fifth determinant, we explored a wide range of R_0_ values, based on a systematic review (Guerra et al., 2017) which reveals a higher variability of measles transmissibility compared to what was previously known.

**Table 1.** Scenarios for evaluating the impact of evidence updates of key determinants of measles vaccination impact. Assumptions for four key determinants used in each scenario and their data sources are summarised. Abbreviations: CFR – case fatality risk, SIA – supplementary immunisation activity.

| Scenarios | Data sources and assumptions for determinants |
| --- | --- |
| Base | With data sources used previously (Verguet et al., 2015; Li et al., 2021):<br>• CFRs: country-specific, time-invariant estimates for those under five years old from Wolfson (Wolfson et al., 2009); assuming CFR is halved after age 5 years.<br>• Contact patterns: physical contact matrix from POLYMOD Great Britain (Mossong et al., 2008).<br>• MCV1 efficacy: step function of age: 85% for under one year old and 95% for over one year old (Sudfeld, Navar & Halsey, 2010).<br>• SIA coverage in zero-dose population: equal coverage in population with or without previous vaccination (random distribution of SIA doses). |
| (A) CFR, Portnoy's method | 'Base' scenario, with updated CFRs: country-specific, time-varying, incidence-related estimates for under and over five years old from Portnoy et al. (Portnoy et al., 2019) |
| (B) Contact patterns, synthetic matrices | 'Base' scenario, with update for contact patterns: country-specific synthetic matrices (Prem, Cook & Jit, 2017; Prem et al., 2020) |
| (B') Contact patterns, proportional mixing | 'Base' scenario, with contact probabilities proportional to the age distribution of population |
| (B'') Contact pattern, uniform mixing | 'Base' scenario, with uniform contact probabilities across age groups |
| (C) MCV1 efficacy, linear age trend | 'Base' scenario, with updated MCV1 efficacy: efficacy as a linear function of age with an increase of 1.49% per month of age, from 68% at birth to 98% at highest. |
| (D) SIA coverage in zero-dose population, dependency on previous vaccination | 'Base' scenario, with update for SIA coverage in zero-dose population: informed by a weighted logistic function fitted to all available surveys (Portnoy et al., 2018) |
| (D') SIA coverage in zero-dose population, dependency on previous vaccination, excluding the largest survey | 'Base' scenario, with update for SIA coverage in zero-dose population: informed by a weighted logistic function fitted to available surveys except for the one with the largest sample size (Indonesia, 2002) (Portnoy et al., 2018) |

**Figure 2.**
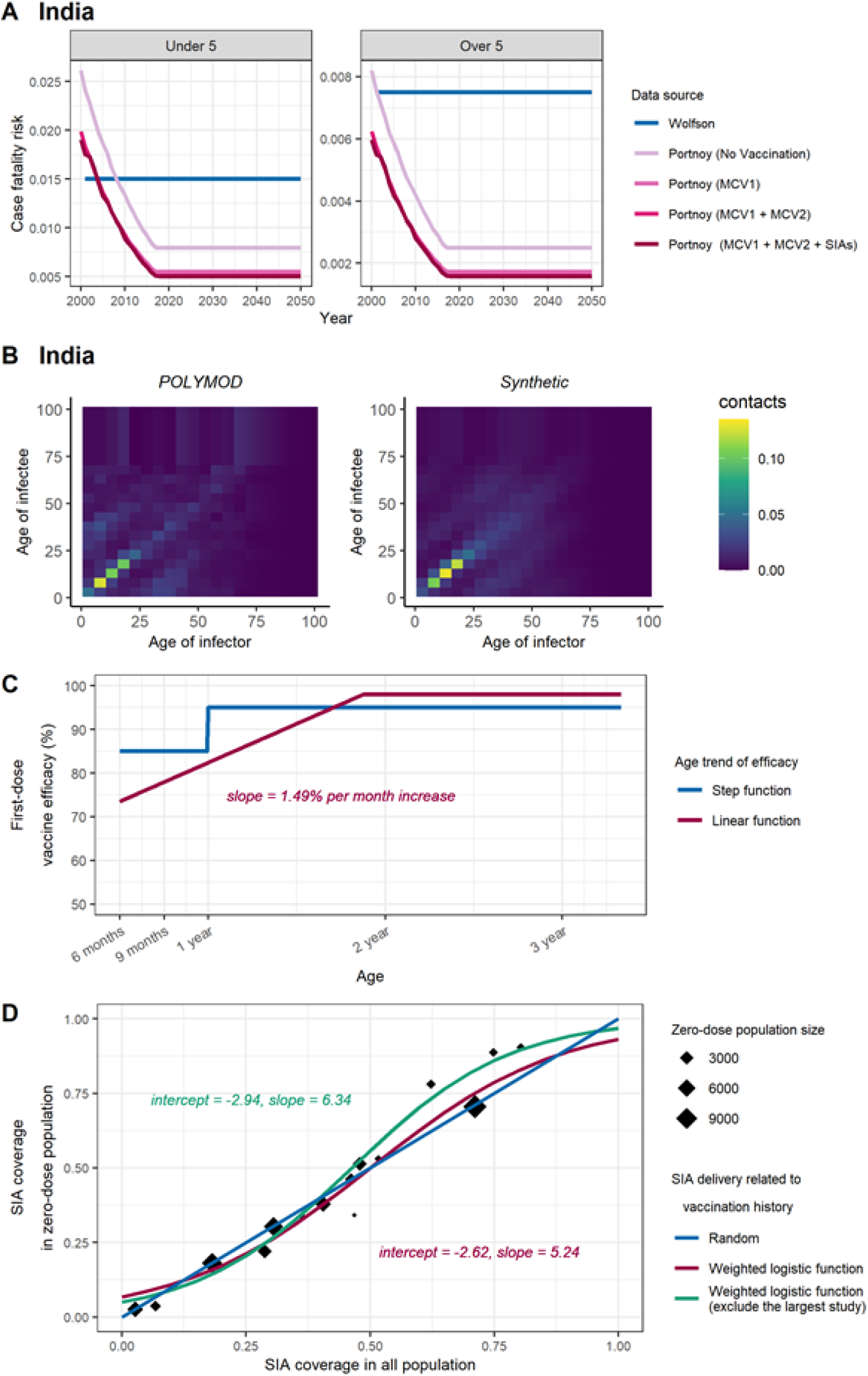
Assumptions for key determinants of measles transmission and vaccination: (A) case fatality risk, (B) social contact patterns, (C) age-dependent vaccine efficacy, and (D) SIA coverage in zero-dose population. Historical and updated data sources for the four determinants are compared. For country-specific assumptions like (A) and (B), we demonstrated the data in India (the country with highest measles mortality in 2000) and include data in other countries in Figure S2 and S3, respectively.

#### (A) Case fatality risk

The impact of measles vaccination using two sets of global CFR estimates was compared (Figure 2A, Figure S2): (i) an earlier review of 58 publications by Wolfson et al. (Wolfson et al., 2009), which provided time-invariant CFRs for children under five years old in different regions, with a further assumption that CFR was halved for children over five years old (Simons et al., 2012); and (ii) an updated review of 124 publications by Portnoy et al. (Portnoy et al., 2019), which estimated country-specific, time-varying CFRs for children under five and over five years old, using a log-linear model with covariates such as local measles attack rate, under-five mortality, calendar year, and percentage of population living in urban areas. For future projection, we assumed that CFRs remained stable at their 2018 levels over 2019-2050, in consideration of uncertain changes in future improvements of healthcare and vaccination programmes as well as the potential impact arising from the COVID-19 pandemic. The Protnoy’s CFR estimates used in this study were also used in a modelling analysis of the impact of COVID-19 disruption to measles vaccination (Gaythorpe et al., 2021).

#### (B) Social contact patterns

Our original study (Verguet et al., 2015; Li et al., 2021) used the contact matrix adapted from the POLYMOD study (Mossong et al., 2008) to represent age-dependent mixing patterns in all countries, because it showed good characterisation of measles transmission. Since then, several contact surveys in LMICs have been conducted, while synthetic contact matrices have been constructed for most countries based on the country-specific household structure, labour force, and school data from LMICs (Prem, Cook & Jit, 2017; Prem et al., 2020). With these two types of social mixing matrices (Figure 2B & FIgure S3), we additionally considered two simple assumptions that are not based on empirical surveys: (i) the POLYMOD contact matrix from Great Britain, (ii) synthetic country-specific contact matrices, (iii) a matrix with contacts proportional to the age distribution of the population, and (iv) a uniform matrix with no age-dependency in mixing (Section S2 in Appendix). All the age-dependent matrices were normalised to reflect the assigned scale of R_0_, so the model results can be properly compared.

#### (C) Age-dependent vaccine efficacy

As shown in Figure 2C, we compared two assumptions for the age trend of first-dose vaccine efficacy: (i) a simplified step-change from 85% to 95% at a cut-off of 1 year old (Sudfeld, Navar & Halsey, 2010); and (ii) a linear increase of 1.49% for every increase in month of age, derived from a recent systematic review of 33 measles vaccine efficacy studies in measles-endemic settings (Hughes et al., 2020). We capped the vaccine efficacy for the first dose at 98%, which is equivalent to the level of protection provided by two doses of MCV.

#### (D) Proportion of zero-dose children reached by SIAs

Based on a previous exploration of the population reached by SIAs using Demographic Health Survey data (Portnoy et al., 2018), we fit a weighted logistic function to describe the association between the SIA coverages in total population and in zero-dose children (Figure 2D). We then calculated the SIA doses given to the population with previous vaccination history (Section S1 in Appendix), by subtracting those received by zero-dose population from the total doses reported in the WHO data (IVB/WHO, 2020a). We compared this association to a random distribution of SIA doses, where SIA doses are delivered to the target population regardless of their measles vaccination history. In addition, to assess the robustness of the weighted logistic function in representing the delivery of SIA, we refitted the function after excluding the survey with the largest sample size (also the oldest survey in the dataset), and then evaluated the vaccination impact.

#### (E) Basic reproduction number

We set R_0_ to 16 in the main analysis, according to a systematic review that included studies in least developed or developing countries in the vaccine era (Guerra et al., 2017). We further examined measles burden and vaccination impact with R_0_ values of 12, 20, and 24, to address the right-skewed distribution of estimates observed in the review (Guerra et al., 2017).

## Results

Figure 3 shows model projections of annual measles incidence over 2000–2050 under different vaccination strategies in India, Nigeria, Pakistan, Ethiopia, and other six high-burden counties in the ‘full-update’ scenario that uses the most recent evidence for four determinants of vaccination impact. Based on coverage projection for MCV1, MCV2, and SIAs during 2000–2050, 252 million measles cases, 3.7 million deaths and 230 million DALYs are projected over the 51 years in the ten countries with high measles burden (Table S3 in Appendix). Among the top ten countries, India contributes the most to overall measles cases (47% with no vaccination) but a smaller proportion of overall deaths (26% with no vaccination), because of its relatively low CFR compared to other countries (Figure S2 in Appendix) (Portnoy et al., 2019).

**Figure 3.**
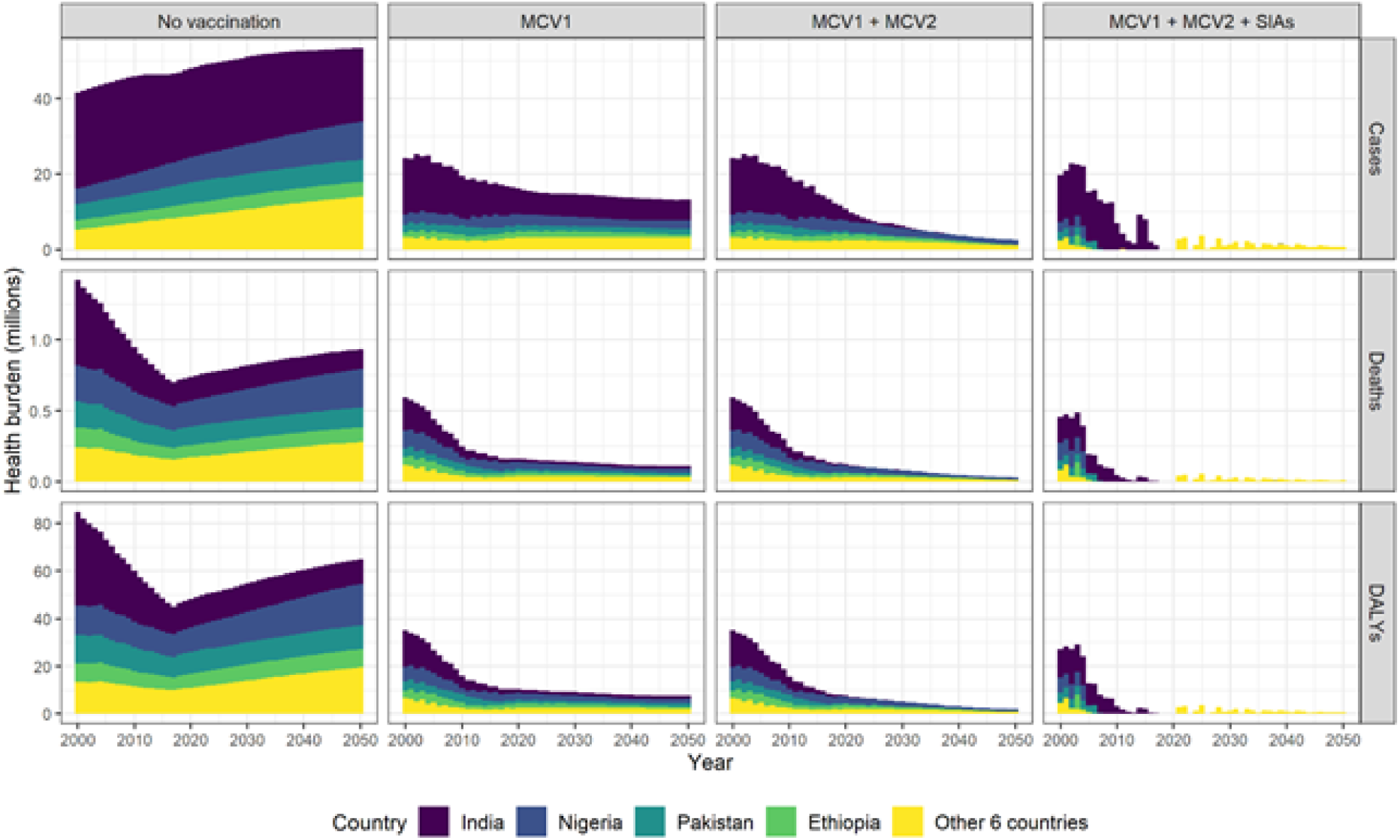
Measles vaccination impact by calendar year. Measles cases, deaths, and disability-adjusted life years by vaccination strategies and countries are presented based on the model results of the ‘full-update’ scenario that includes all the updates of key determinants. In India, Nigeria, Pakistan, Ethiopia, and other 6 countries (Afghanistan, Sudan, Tanzania, Niger, Somalia, DR Congo) with high measles burden, a substantial reduction of measles burden can be attributed to MCV1, while MCV2 and SIAs contribute to the maintenance of low-level measles transmission.

Model results show the important roles of continuous routine and SIA vaccination delivery in measles control. Of these vaccination activities, the first dose provided through routine programmes (MCV1) contributes the most to burden reduction. Based on the ‘full-update’ model, cumulative measles cases over 2000–2050 are projected to reduce by 66% with MCV1 alone, 73% with a second routine dose (MCV2), and 89% with further inclusion of SIAs. The incremental impact of MCV2 and SIA doses is not as large as MCV1 but essential in maintaining a low level of measles transmission and preventing the accumulation of susceptibles that can lead to large sporadic outbreaks. With continuous improvement in MCV1, MCV2, and SIAs, measles incidence is projected to remain <1 case per million for three consecutive years in seven out of the ten high-burden countries during the projection period (Table S2 in Appendix). Conversely, Nigeria, Somalia, and DR Congo are not likely to maintain this low level of measles incidence prior to 2050, as outbreaks are projected every few years.

Table 2 presents model projections of vaccination impact in terms of total averted cases, deaths, and DALYs over 2000–2050, under alternative scenarios for updated evidence sources informing CFR, contact pattern, age-dependent vaccine efficacy, and SIA delivery to zero-dose children. Measles burden for each country, vaccination strategy, and scenario for evidence update is included in Table S2 in Appendix. Using the updated CFR estimates from Portnoy’s review instead of Wolfson’s review is the most influential single change, resulting in 17.0–22.9% reduction in averted deaths and DALYs. Using contact patterns from synthetic matrices or the POLYMOD Great Britain matrix results in similar estimates of vaccine-averted cases and deaths, lying within 3% of each other. Under the assumptions of proportional and uniform mixing patterns, vaccine-averted cases also remain similar to the estimates based on the POLYMOD matrix. However, the assumptions of uniform mixing and proportional mixing result in a 22.9–26.0% and 7.3–99.5% reduction in the number of averted deaths respectively due to the increase in the mean age of measles infection and death (Figure 4). Next, applying a linear trend in vaccine efficacy by age instead of a step function results in a 2.0-9.5% decline in averted cases, depending on the vaccination strategy. Finally, the inclusion of a weak dependency between SIA doses and previous vaccination is projected to only produce a small reduction in averted cases (0.17–0.27%), using either of the logistic functions derived from previous surveys, as the delivery of SIA doses to the target population is nearly random (Figure 2D). Overall, the evidence updates about the CFR estimates and age trend in vaccine efficacy have the largest influence on averted health burden of measles.

**Table 2.** Averted measles cases, deaths, and DALYs by vaccination strategy and evaluation scenario. Total vaccine-averted burden in ten high measles burden countries over 2000–2050 is presented in a unit of millions. Percentages in parentheses are the proportionate change from ‘base’ scenario. All model scenarios were conducted under the assumption of R_0_ = 16. Details of data sources for key determinants assumed for each scenario can be found in Table 1. Abbreviations: CFR – case fatality risk, MCV1 – measles containing vaccine, first dose, MCV2 – measles containing vaccine, second dose, SIA – supplementary immunisation activity.

| Scenarios | MCV1 |  |  | MCV1 + MCV2 |  |  | MCV1 + MCV2 + SIAs |  |  |
| --- | --- | --- | --- | --- | --- | --- | --- | --- | --- |
|  | Cases, millions | Deaths, millions | DALYs, millions | Cases, millions | Deaths, millions | DALYs, millions | Cases, millions | Deaths, millions | DALYs, millions |
| Base | 1799<br>(ref) | 47<br>(ref) | 3149<br>(ref) | 2017<br>(ref) | 51<br>(ref) | 3387<br>(ref) | 2259<br>(ref) | 58<br>(ref) | 3823<br>(ref) |
| (A) CFR, Portnoy’s method | - | 39<br>(-17.0%) | 2585<br>(-17.9%) | - | 41<br>(-19.6%) | 2693<br>(-20.5%) | - | 45<br>(-22.3%) | 2946<br>(-22.9%) |
| (B) Contact pattern, synthetic matrices | 1820<br>(1.2%) | 46<br>(-1.8%) | 3094<br>(-1.7%) | 2018<br>(0.05%) | 49<br>(-2.9%) | 3288<br>(-2.9%) | 2269<br>(0.43%) | 56<br>(-3.1%) | 3704<br>(-3.1%) |
| (B’) Contact pattern, proportional mixing | 1811<br>(0.72%) | 44<br>(-7.3%) | 2909<br>(-7.6%) | 1977<br>(-2.0%) | 46<br>(-8.8%) | 3068<br>(-9.4%) | 2218<br>(-1.8%) | 53<br>(-8.9%) | 3459<br>(-9.5%) |
| (B’’) Contact pattern, uniform mixing | 1810<br>(0.65%) | 36<br>(-22.9%) | 2410<br>(-23.5%) | 1960<br>(-2.8%) | 38<br>(-24.6%) | 2522<br>(-25.5%) | 2228<br>(-1.4%) | 44<br>(-24.6%) | 2830<br>(-26.0%) |
| (C) First-dose vaccine efficacy, linear age trend | 1612<br>(-10.4%) | 43<br>(-9.6%) | 2848<br>(-9.6%) | 1932<br>(-4.3%) | 48<br>(-4.9%) | 3229<br>(-4.7%) | 2216<br>(-1.9%) | 57<br>(-2.1%) | 3747<br>(-2.0%) |
| (D) SIA delivery to zero-dose population, dependency on previous vaccination | - | - | - | - | - | - | 2265<br>(0.27%) | 58<br>(0.18%) | 3830<br>(0.18%) |
| (D’) SIA delivery to zero-dose population, dependency on previous vaccination | - | - | - | - | - | - | 2263<br>(0.17%) | 58<br>(0.16%) | 3829<br>(0.15%) |
| Full-update | 1643<br>(-8.7%) | 36<br>(-24.2%) | 2364<br>(-24.9%) | 1942<br>(-3.7%) | 38<br>(-25.8%) | 2512<br>(-25.8%) | 2245<br>(0.62%) | 43<br>(-26.2%) | 2801<br>(-26.7%) |

**Figure 4.**
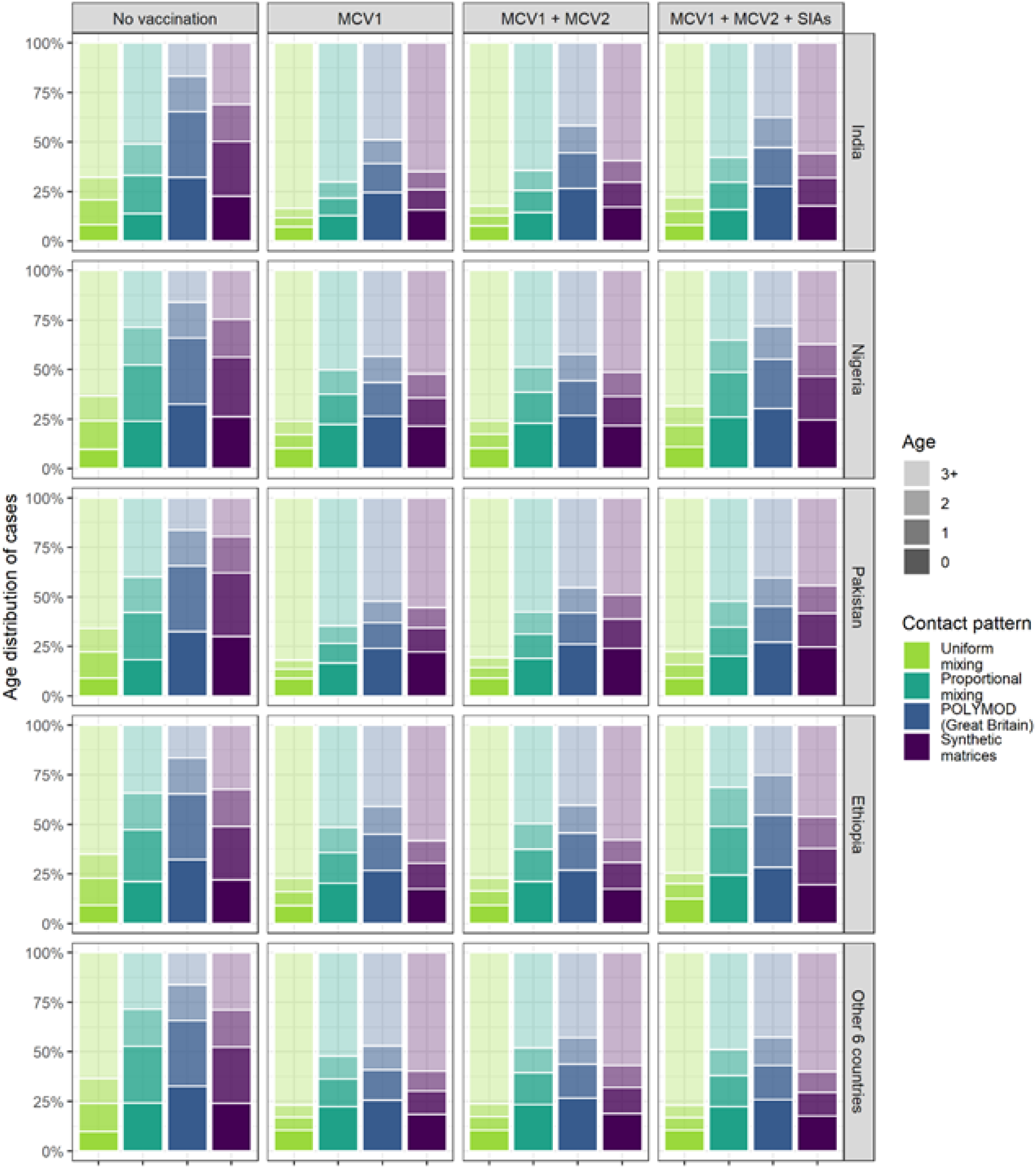
Age distribution of cumulative measles cases over 2000–2050 by different contact patterns. Proportions of measles cases aged 0, 1, 2, and 3+ years in India, Nigeria, Pakistan, Ethiopia, and other 6 countries (Afghanistan, Sudan, Tanzania, Niger, Somalia, DR Congo) with high measles burden are presented, by the assumptions of uniform mixing, proportional mixing, POLYMOD Great Britain contact matrix, and country-specific synthetic contact matrices. Compared to uniform mixing, contact patterns that consider age-related mixing tend to result in a lower average age at infection. Based on the POLYMOD matrix, the projected cases are more concentrated in younger age groups.

The combined effect of all these evidence updates on the key determinants of measles vaccination impact is a decline in vaccine-averted burden, with 0.62–8.7% decrease in cases averted, and 24.2– 26.7% decrease in deaths and DALYs averted by measles vaccination. However, the relative impact of MCV1, MCV2, and SIA doses to measles burden reductions do not change with these evidence updates.

Next, we compare the age distribution of cumulative measles cases over 2020-2025 under different assumptions for social contact patterns. Figure 4 presents the proportions of measles cases from zero, one, two, and three years and above of age in India, Nigeria, Pakistan, Ethiopia, and other six high-burden countries. Similar age distributions of cases are shown across countries. The update with country-specific synthetic matrices results in an older average age of measles cases, compared to the POLYMOD matrix. The age distribution of measles cases is least concentrated in younger age groups under the assumption of uniform mixing. Without the implementation of vaccination, the proportions of cases older than three years old in each country are 16–17% with the POLYMOD matrix, 20–32% with synthetic matrices, 22–51% with proportional mixing, and 62–68% with uniform mixing. The increase in the mean age under the assumption of uniform mixing accounts for the reduced vaccination impact against deaths, since older measles cases are at a lower risk of death (Figure S2), even though there are more cases prevented by vaccination (Table 1). We also observe a shift towards an older average age at infection in any of the measles vaccination strategies, compared to the model results assuming no vaccination.

Overall, there are no apparent synergistic or antagonistic effects of the vaccination impact from the combined updates of the first four determinants discussed in this study; that is, the total change in vaccine-averted burden is approximately the sum of changes from individual determinants. Under the DynaMICE model structure of measles transmission and vaccination, there was no strong interaction between the key determinants that could largely modify measles vaccination impact.

Figure 5 presents the absolute measles burden and vaccination impact over the evaluation period by different values of R_0_ in the ‘full-update’ scenario. A larger value of R_0_ led to a higher estimate of measles burden but a smaller vaccination impact. However, the overall impact of changing R_0_ is limited, with less than 5% of variation in cases and deaths averted by vaccination. Increased burden driven by a larger R_0_ also delays the achievement of measles elimination, in particular for countries with wider gaps in vaccine coverage (Table S2 in Appendix). Nevertheless, our findings about the substantial contribution of measles vaccination and the relative impact of MCV1, MCV2 and SIA doses remain unchanged.

**Figure 5.**
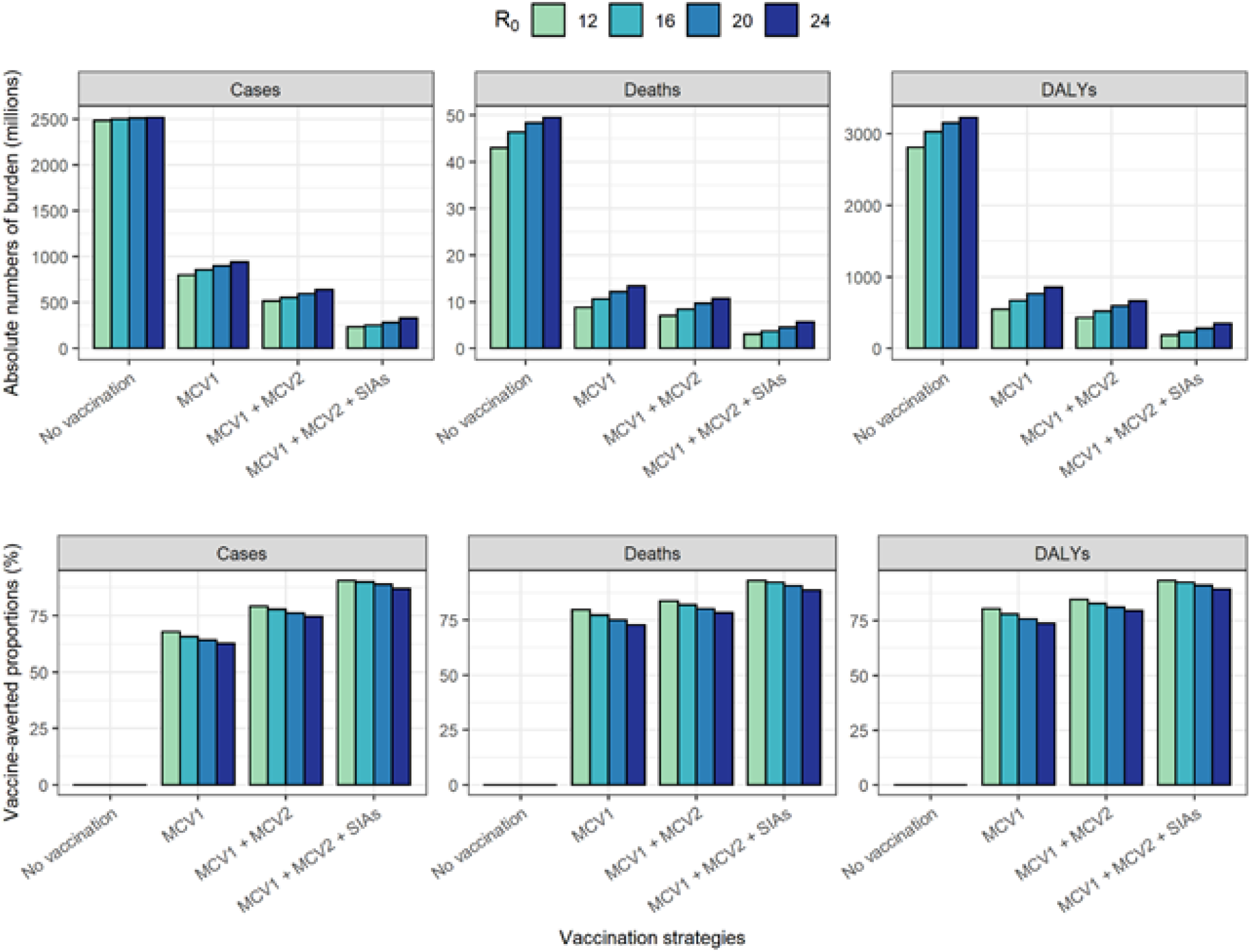
Cumulative measles burden over 2000–2050 by vaccination strategies and R_0_. The total number of cases, deaths, and DALYs (upper panel) and corresponding averted proportions (lower panel) in the four countries are compared by different values of R_0_. Despite the variability of R_0_, there is a limited effect on the vaccine impact estimates.

## Discussion

Using the DynaMICE model, we have considered the best available updated evidence and explored the influence of recent systematic reviews and database analyses on key determinants of measles vaccination impact: case fatality risk, social contact patterns, age-dependent vaccine efficacy, proportion of zero-dose children reached by SIAs, and basic reproduction numbers. Despite large changes in many of these underlying parameters, we find that our previous findings about measles burden and vaccination impact still broadly hold. As before, we find that the first vaccine dose delivered by routine immunisation programmes contributes to the largest reduction of measles burden, but the second routine dose and supplementary doses also provide important incremental benefits towards elimination. We find that updates to two of the determinants have the biggest changes in the vaccination impact: making the first-dose vaccine efficacy a linearly age-dependent function based on a recent review (Hughes et al., 2020) decreases impact estimates by 2–10%, while using time-dependent and incidence-related CFRs (Portnoy et al., 2019) decreases them by 17–23%. Updated contact patterns, SIA dependency on previous vaccination history and the basic reproduction number make relatively little difference to vaccine impact estimates. Overall, measles vaccination impact remains very substantial despite these data updates.

In the ten countries with the highest measles mortality in 2000, our model projections show varying trends of measles incidence under the vaccination strategy involving MCV1, MCV2 and SIAs doses. In Nigeria, Somalia, and DR Congo, measles transmission is projected to be suppressed for a few years followed by resurgence (Table S2 in Appendix). The difficulties faced by these countries result from having a lower coverage of routine immunisation (both MCV1 and MCV2) compared to the other countries, and their SIAs are not highly effective to close the measles immunity gaps (Figure S1 in Appendix). We did not model the possibility of measles elimination directly, since this would require consideration of phenomena such as case importation, contact tracing and outbreak response that are not captured by aggregate compartmental models such as DynaMICE. However, the low measles incidence achieved by countries like India suggest that the measles elimination schedules aimed by the WHO regional offices (WHO, 2019); WHO SEAR, 2020) may be possible if high coverage of both routine and campaign vaccination activities can be maintained (Table S2, Figure S4 & Figure S5 in Appendix).

Updating CFR estimates is the single most influential change among the determinants of measles vaccination impact that we evaluated. The updated Portnoy’s CFRs start higher than Wolfson’s CFRs in 2000 but then rapidly decline (Figure S2 in Appendix), whereas Wolfson’s CFRs are assumed to be time-invariant. In Portnoy’s method, higher measles CFR is associated with higher incidence, but it is possible that any large pressure on the healthcare system could be reflected in CFR estimates. Across the ten high-burden countries, a 17–22% reduction is seen in cumulative vaccine-averted deaths and DALYs over 2000–2050 with updated CFRs. Additionally, with more doses included in the vaccination strategy, averted measles burden is projected to increase as a result of a larger decline in Portnoy’s CFRs (Figure S2 in Appendix). Referring to the log-linear model used for estimating CFRs (Portnoy et al., 2019), we found measles attack rate, one of the covariates which can be largely reduced by vaccination, may explain the different levels of reduction in vaccination impact across vaccination strategies. Given the uncertainty about future changes in the determinants of measles mortality (such as nutrition and healthcare access), particularly in the wake of societal disruption caused by the COVID-19 pandemic, we assume that no further improvements to CFRs occur beyond 2018. Considering the importance of CFRs to estimates of measles burden and vaccination impact, understanding how CFRs may change in the future needs to be a key future research priority.

The relationship between MCV1 efficacy and vaccination age is another important determinant of vaccination impact. In our updated analysis, we assumed that MCV1 efficacy varies linearly with age (Hughes et al., 2020), instead of being constant when given at any age below 1 year (Sudfeld, Navar & Halsey, 2010) (Figure 2C). Since the measles vaccine is mostly given to young children, this reduces our estimates of MCV1 efficacy and hence of MCV1 impact (Table 2). However, SIA doses (which reach wide target age ranges mostly above the age at which MCV1 is given) may show increased impact with this update (Table 2).

Like previous modelling studies (Wallinga et al., 2001; Funk et al., 2019), our analysis shows the importance of age-dependent contact patterns in determining measles transmission and the impact of control strategies. Despite a small difference in the vaccination impact on preventing measles cases, a 23–25% reduction in deaths averted is projected under the assumption of uniform mixing, compared to the POLYMOD matrix (Table 1), due to the increase in the average age at measles infection (Figure 3). Compared to uniform mixing, proportional mixing also relates to measles cases of a younger age, reflecting the age composition of local demographics that consists of a larger proportion of children in most high measles burden settings. On the other hand, there is little difference in model results between using the POLYMOD Great Britain matrix (Mossong et al., 2008) and country-specific synthetic matrices (Prem et al., 2020). Both matrices capture the main pattern of age-assortative mixing and thus led to similar estimates of vaccination impact. The slight difference in age distribution of measles cases could be attributed to the higher degree of age-assortativity in the POLYMOD Great Britain matrix (Figure S2 in Appendix), which might be less representative of low-and middle-income settings with more frequent contact between different age groups.

Vaccination impact is least affected by including the dependency of SIA delivery on previous history of measles vaccination (<0.3%) (Table 2). Relying on the updated data (Portnoy et al., 2018), we fitted logistic function indicates SIAs are more likely than average to reach zero-dose children when the coverage in the total population is between 50–85% (Figure 2D), despite a weak association.

However, those surveys used for the evidence update were conducted more than a decade ago (Portnoy et al., 2018), and more recent SIAs may have been better able to address subnational inequalities in vaccination coverage, e.g., by targeting rural areas or hard-to-reach populations (Local Burden of Disease Vaccine Coverage Collaborators, 2020).

Our results present a substantial decline in measles cases and deaths over 2000–2019 in ten high-burden countries. Such decline is also seen in the GBD estimates (GBD 2019 Diseases and Injuries Collaborators, 2020), although our results show relatively lower burden estimates (Figure S4 in Appendix). In the GBD estimates, the country rank of measles mortality in 2019 has changed and India is no longer the country with the highest burden. This difference in the country-level burden is consistent with our findings (Table S2 in Appendix), reflecting the variations in the MCV1, MCV2, and SIA coverage. Additionally, we also compared the model outputs to cases collected from local measles surveillance networks (IVB/WHO, 2020b). We particularly noticed that in Pakistan, Sudan, and Niger, our model estimates suggested that country-specific measles incidence is maintained below 1 per million prior to 2020, with the implementation of both routine and SIA vaccination (Figure S4 in Appendix), while there were more than 2,000 cases notified to the WHO surveillance system in 2019. The inconsistency between model estimates and reported data may be associated with outbreaks related to heterogeneous vaccine coverage at the sub-national level (Mere et al., 2019; Local Burden of Disease Vaccine Coverage Collaborators, 2020), which are not precisely captured by a model based on national averages of vaccine coverage. However, underreporting in measles surveillance data complicates any direct comparisons between model estimates and notified cases (Simons et al., 2012).

Our analysis has some limitations. First, we assumed that MCV1 is delivered promptly to children at 9 months old, which assumes perfect adherence to the routine immunisation schedule. This assumption of perfect timeliness may not be realistic in most settings, where many children receive MCV1 earlier, or more likely, later than the targeted age (Clark & Sanderson, 2009). Thus, setting-specific data on the precise age at vaccination could provide further insight into vaccination impact. Additionally, we assumed the same R_0_ across countries due to a lack of systematic data for estimating the country-specific transmissibility. Combining with the age-dependent contact patterns, measles models fitted to data on incidence and serology may be helpful to capture the force of measles infection in each study country (Funk et al., 2019). Nonetheless, our model findings remain robust and useful, as we found in sensitivity analyses that variations in R_0_ within the range 12–24 do not substantially affect the vaccination impact (Figure 4). Third, we did not consider importation of measles cases from migration, which can be a major source of local outbreaks when domestic measles transmission has been largely eliminated (Ramsay et al., 2003). Fourthly, we assumed that vaccine coverage was uniform over the entire country. However, the countries we modelled all have subnational regions with substantially lower coverage than average, and which therefore sustain measles transmission even when it is eliminated in the rest of the country.

Recent advances in data and research have provided better understanding of measles transmission and vaccination impact. Using the DynaMICE model, we have systematically assessed updates on the key determinants of measles epidemiology and vaccination in ten countries with historical high measles mortality. We identified that measles vaccination impact is sensitive to the assumptions on CFR and age-dependent vaccine efficacy and reassure the importance of age-dependent social contact structure. While we recommend incorporating these evidence updates into measles models in the future analysis, uncertainty in the collected evidence and how they are translated into model inputs should not be overlooked. As more evidence on the key determinants of vaccination impacts are available, an updated review and examination on the process of data synthesis will be required. However, as we have demonstrated in this study, these evidence updates would not undermine the substantial contribution of measles vaccination. High coverage of both measles vaccine doses, either through routine or SIA vaccine delivery, are essential for meeting and maintaining the goals for measles elimination.

## Data Availability

Country-specific coverage data for measles vaccination were extracted from public databases of the World Health Organization. Population statistics were obtained from the United Nations World Prospect Project 2019.

https://apps.who.int/immunization_monitoring/globalsummary/timeseries/tswucoveragemcv1.html

https://apps.who.int/immunization_monitoring/globalsummary/timeseries/tswucoveragemcv2.html

https://www.who.int/entity/immunization/monitoring_surveillance/data/Summary_Measles_SIAs.xls

https://population.un.org/wpp/

## Competing interests

The authors have declared that no competing interests exist.

## Acknowledgements

We would like to thank Dr Kiesha Prem for providing helpful guidance on the application of synthetic contact matrices in this modelling analysis.

This work was carried out as part of the Vaccine Impact Modelling Consortium (www.vaccineimpact.org), but the views expressed are those of the authors and not necessarily those of the Consortium or its funders. The funders were given the opportunity to review this paper prior to publication, but the final decision on the content of the publication was taken by the authors.

This work was supported, in whole or in part, by the Bill & Melinda Gates Foundation, via the Vaccine Impact Modelling Consortium [Grant Number INV-009125]. Under the grant conditions of the Foundation, a Creative Commons Attribution 4.0 Generic License has already been assigned to the Author Accepted Manuscript version that might arise from this submission.

## Notes

### Competing Interest Statement

The authors have declared no competing interest.

### Author Declarations

This modelling analysis was based on publicly available data. No individual patient data were used.

